# Clinical and laboratory characteristics in outpatient diagnosis of COVID-19 in healthcare professionals in Rio de Janeiro, Brazil

**DOI:** 10.1101/2020.10.22.20217851

**Authors:** Luís Cristóvão Porto, Claudia Henrique da Costa, Alessandra Sant’Anna Nunes, Isabel Bouzas, Tiago C. Ferreira, Vinicius M. Porto, Danielle A. Secco, Sandra P. C. Vilas Boas, Anamelia C. Faria, Rosangela A. G. Marins, Cíntia A. Duarte, Elizabeth P. Bittencourt, Anderson V. Loureiro, Angela Maria G. Santos, Vania Maria A. Souza, Fernanda H. Silva, Maria Cristina Lopes, Marcelle V. Castro, Areta D. Silva, Flavia M. Vasconcelos, Diego S. Moreira, Juliana P. Motta, Cristiano V. Lima, Raphaela S. Menezes, Nathalia B. V. Brazão, Jeane S. Nogueira, Ana Paula Villela Silva, Ingrid A.A. Brito, Scarlathe B. Costa, Roberta Mariana R. Lemes, Jessica Isis O. dePaula, Giovanna L. Bongionanni, Ohana C. L. Bezerra, Valter V. Andrade-Neto, Leonardo Morette, Rafael S. Barbosa, Jose Afonso L. Sanches, Pedro L. Melo, Sergio M. Freire, Romulo Souza, Laiz Gomes, Denize P. Felizardo, Alexsandre V. Rodrigues, Priscila A. Franco, Renata Miranda, Rogerio Rufino

**Affiliations:** Histocompatibility and Cryopreservation Laboratory, Institute of Biology Roberto Alcantara Gomes, Rio de Janeiro State University, Rio de Janeiro, 20950-003; Clinical Pathology Service, Polyclinic Piquet Carneiro, Rio de Janeiro State University, Rio de Janeiro, 20950-003; Department of Pulmonology and Tisiology, Faculty of Medical Sciences and Service of Pulmonology, Polyclinic Piquet Carneiro, Rio de Janeiro State University, Rio de Janeiro, 20950-003; Nurse Department, Polyclinic Piquet Carneiro, Rio de Janeiro State University, Rio de Janeiro, 20950-003; Health Research Support Facility Center (CAPCS), Polyclinic Piquet Carneiro, Rio de Janeiro State University, Rio de Janeiro, 20950-003; Infectology Service, Polyclinic Piquet Carneiro, Rio de Janeiro State University, Rio de Janeiro, 20950-003; Research Division, Clementino Fraga Filho University Hospital, Federal University of Rio de Janeiro, Rio de Janeiro, 21941-590; Administrative and Finance Services, Polyclinic Piquet Carneiro, Rio de Janeiro State University, Rio de Janeiro, 20950-003; Pharmacy and Supply Service, Polyclinic Piquet Carneiro, Rio de Janeiro State University, Rio de Janeiro, 20950-003; Logistic, Informatic and Infrastructure Service, Polyclinic Piquet Carneiro, Rio de Janeiro State University, Rio de Janeiro, 20950-003; Laboratory of Biomedical Instrumentation, Institute of Biology Roberto Alcantara Gomes, Rio de Janeiro State University, Rio de Janeiro, 20550-013; Information Technology and Education in Health Department, Faculty of Medical Sciences, Rio de Janeiro State University, Rio de Janeiro, 20551-031; Communication and Human Resource Service, Polyclinic Piquet Carneiro, Rio de Janeiro State University, Rio de Janeiro, 20950-003

**Keywords:** diagnosis, leukocytes, biochemistry, COVID-19, healthcare professionals, leukocytes, anosmia, body pain, C-reactive protein, monocytes, fever, chills

## Abstract

**Aims:** This study aimed to identify the symptoms associated with early stage SARS-CoV-2 (COVID-19) infections in healthcare professionals (HCP) using both clinical and laboratory data.

**Methods:** A total of 1,297 patients, admitted between March 18 and April 8, 2020, were stratified according to their risk of developing COVID-19 using their responses to a questionnaire designed to evaluate symptoms and risk conditions.

**Results:** Anosmia/hyposmia (p <0.0001), fever (p<0.0001), body pain (p<0.0001), and chills (p=0.001) were all independent predictors for COVID-19, with a 72% estimated probability for detecting COVID-19 in nasopharyngeal swab samples. Leukopenia, relative monocytosis, decreased eosinophil values, CRP, and platelets were also shown to be significant independent predictors for COVID-19.

**Conclusions:** The significant clinical features for COVID-19 were identified as anosmia, fever, chills, and body pain. Elevated CRP, leukocytes under 5,400 x 10^9^/L, and relative monocytosis (>9%) were common among patients with a confirmed COVID-19 diagnosis. These variables may help, in the absence of RT-PCR tests, to identify possible COVID-19 infections during pandemic outbreaks.

**Summary:** From March 19 to April 8 2020, 1,297 patients attended the Polyclinic Piquet Carneiro for COVID-19 detection. Healthcare professional data was analyzed, significant clinical features were anosmia, fever, chills and body pain. Elevated CRP, leukopenia and monocytosis were common in COVID-19.

## INTRODUCTION

The coronavirus disease 2019 (COVID-19) pandemic caused by the outbreak of the novel severe acute respiratory syndrome coronavirus-2 (SARS-CoV-2) represents a significant and urgent threat to global health. By October 6, 2020, Brazil ranks third in the number of COVID-19 cases (4,927,235) and second in number of deaths (146,675) in the world {University, 2020 #1800}. Outbreaks of this magnitude lead to significant increases in demand for hospital beds, medical equipment, and human resources. Additionally, healthcare professionals (HCP) working on the frontline are amongst the groups most affected by the pandemic - in Italy, for example, 20% of these HCP were affected by COVID-19,[2] contributing to an even greater burden on the health system. Despite public health responses aimed at containing the disease and delaying its spread, several countries have been challenged with an intensive care crisis, and this crisis is imminent in Brazil. In this context, attention to the health and wellbeing of HCP is essential to maintain the integrity and functionality of the health system which allows for both population care and pandemic surveillance.

In this study, we investigated the clinical features of 1,297 individuals, mostly HCP (80%), associated with the presence of the SARS-CoV-2 (COVID-19) during the period immediately following the declaration of the pandemic in Brazil.

## MATERIALS AND METHODS

### Patient selection and characteristics

A total of 1,297 individuals visited the Piquet Carneiro Polyclinic at the Rio de Janeiro State University (UERJ), a designated diagnostic site for personnel working in the Brazilian public health system, for screening of COVID-19 between March 19 and April 8, 2020. Here, they found three levels of assistance. First, both HCP and clinical patients were asked to identify their general symptoms. Following this, asymptomatic patients and HCP who had appointments scheduled at various specialized clinics were provided with a white bracelet and allowed to enter the medical center for their consultation. Then, symptomatic patients were referred to the care of the nursing and medical teams. All patients with dyspnea, high fever, chest pain, abnormal pulmonary auscultation were then prescribed chest ultrasound to identify if hospital admission was necessary. Individuals were then immediately assigned to various clinical grades in accordance with those patients who presented with a fever (>37.5°C) or poor O_2_ saturation (<94%) and were assigned yellow or red bracelets. After this evaluation, patients with red bracelets were directed to the hospital when necessary and the rest of the patients were asked to complete the interview questionnaire. They received the necessary medical assistance and laboratory samples were collected. All patients with yellow or red bracelets received a full clinical and laboratory screening.

The questionnaire was used to identify high-risk comorbidities, influenza vaccine status, and to facilitate the contact tracing over the previous two weeks (Table 1). Signs and symptoms, nasopharyngeal swabs (NPS), and blood samples for the complementary biochemistry (n=1,297) and hematology tests (n=1,211) were also collected from all the patients. This study was approved by the Local and National Ethics Committee (CAAE: 30135320.0.0000.5259).

**Table 1.** Characteristics of individuals attended for COVID-19 detection from March 18 to April 8, 2020 at Polyclinic Piquet Carneiro, Rio de Janeiro

|  | HCP | Non-HCP | Total | P |
| --- | --- | --- | --- | --- |
| N | **1,001** | **266** | **1,297** |  |
| Female n (%) | 755 (58.2) | 155 (75.4) | 910 (71.8) | **<0.0001** |
| Age (x ± SD) | 41.3 ± 10.6 | 42.4 ± 11.8 |  | 0.956 |
| BMI median [25-75] | 26.5 [23.8-30.4] | 28.3 [24.7-33.2] |  | **0.0009** |
| **Color-Race n (%)** | **935** | **244** | **1,179** |  |
| White | 532 (56.9) | 99 (40.6) | 631 (53.5) | **0.0004** |
| Brown | 248 (26.5) | 85 (34.8) | 333 (28.2) |  |
| Black | 135 (14.4) | 55 (22.5) | 190 (16.1) |  |
| Asian | 11 (1.2) | 3 (0.1) | 14 (1.2) |  |
| Not-declared | 8 (0.8) | 2 (0.1) | 10 (0.8) |  |
| Indigenous | 1 (0.0) | 0 (0) | 1 (0.0) |  |
| **Education n (%)** | **953** | **243** | **1,369** |  |
| Less 9y | 3 (3.1) | 36 (14.8) | 39 (3.3) | **<0.0001** |
| 9-12y | 185 (19.4) | 92 (37.9) | 277 (23.2) |  |
| 12-16y | 355 (37.2) | 85 (35.0) | 440 (36.8) |  |
| >16y | 410 (43.0) | 30 (12.3) | 440 (36.8) |  |
| **Vaccine status** |  |  |  |  |
| Influenza vaccine in 2019 | 636 (76.0) | 114 (61.9) | 750 (73.5) | **<0.0001** |
| Influenza vaccine in 2020 | 375 (47.7) | 53 (31.1) | 428 (44.8) | **<0.0001** |
| **Medical History n (%)** | **1,001** | **266** | **1,297** |  |
| Arterial Hypertension | 180 (17.9) | 65 (24.4) | 245 (19.3) | **0.012** |
| Medicine drugs (in use) | 154 (15.3) | 28 (10.5) | 182 (14.3) | **0.025** |
| Chronic Sinusitis | 81 (8.0) | 36 (13.5) | 117 (9.2) | **0.006** |
| Asthma | 86 (8.5) | 18 (6.7) | 104 (8.2) | 0.203 |
| Contraceptives (n=1052) | 78 (10.3) | 22 (14.1) | 100 (10.9) | 0.106 |
| Diabetes | 58 (5.7) | 20 (7.5) | 78 (6.1) | 0.184 |
| Tobacco use | 44 (4.4) | 29 (10.9) | 73 (5.7) | **<0.0001** |
| Obesity | 42 (4.2) | 30 (11.2) | 72 (5.6) | **<0.0001** |
| Chronic bronchitis | 50 (5.0) | 14 (5.2) | 64 (5.0) | 0.481 |
| Alcohol addiction | 47 (4.7) | 7 (2.6) | 54 (4.2) | 0.090 |
| Hypothyroidism | 43 (4.3) | 3 (1.1) | 46 (3.6) | **0.007** |
| Cardiovascular disease | 19 (1.9) | 6 (2.2) | 25 (1.9) | 0.432 |
| Tuberculosis | 10 (1.0) | 4 (1.5) | 14 (1.1) | 0.336 |
| Mental diseaseL | 4 (0.4) | 4 (1.5) | 8 (0.6) | 0.065 |
| Neoplasia | 5 (0.5) | 1 (0.3) | 6 (0.4) | 0.631 |
| Liver Disease | 3 (0.3) | 3 (1.1) | 6 (0.4) | 0.111 |
| Rheumatoid Arthritis | 5 (0.5) | 0 (0) | 5 (0.3) | 0.307 |

HCP – Healthcare professionals, Non-HCP – non-Healthcare personnel. BMI – body mass index. Data <expressed as mean+/- SD, compared with ANOVA or median [25%-75%].

### Real-time PCR SARS-COV-2 detection and laboratory testing

RNA was extracted from the samples according to both the COVID-19 infection and laboratory detection for the SARS-COV-2 guidelines [12]. Specific real-time reverse transcription polymerase chain reaction (RT-PCR) assays were performed using commercial kits or previously designed nCoVqRT-PCR kits (Biomanguinhos, Fiocruz, Rio de Janeiro, and Instituto de Biologia Molecular do Paraná, Paraná) approved by the Brazilian Vigilance (ANVISA) group within the Histocompatibility and Cryopreservation Laboratory at UERJ.

Peripheral blood tests were performed by the Pathology Service of the Piquet Carneiro Polyclinic using several automated systems. The complete blood counts were carried out using a SYSMEX XT1800i, while the C-reactive protein (CRP; immunoturbidimetric), creatinine (CR, picrate kinetic), lactate dehydrogenase (LDH; IFCC), aspartate and alanine aminotransferases (AST and ALT; NAPH), and creatine phosphokinase (CK; N-acetyl L-cysteine) assays were performed using the Architect c8000 Equipment from Abbott.

### Data collection

All the questionnaire, nursing and medical consultation records, and laboratory findings were added to the electronic media files for each patient. This data was then used to ascertain the epidemiological and symptom data for the larger cohort. All data were checked by two independent physicians (LCP and IB).

### Statistical analysis

The quantitative peripheral blood tests were compared using ANOVA or the Mann-Whitney U test and the Kruskal Wallis tests when appropriate. The categorical variables were expressed as numbers (%) and compared using the Fisher’s exact test. Differences were considered significant at a p-value of < 0.05 using a one-tailed test. Tukey or Dunn test was applied when multiple comparisons were analyzed, respectively for normal or nonparametric variables. The date of testing was also grouped by week. In the multivariate analyses, Binary Logistic Regression (LR) was used to identify the clinical, laboratory and independent signs or symptoms that could explain (or predict) the presence of the SARS-COV-2 in each sample. The selection process was applied using a stepwise process which allowed us to select the smallest subgroup of independent variables that best explained any particular outcome. All analyses were performed using the SPSS 26.0 software.

## RESULTS

### Demographic and laboratory characteristics linked to positive COVID-19 test results

Risk factors related to the COVID-19 NPS results are summarized in Tables 2 and 3.

**Table 2.** Medical history and vaccine status from COVID-19 nasopharyngeal swab individuals attended from March 18 to April 8, 2020 at Polyclinic Piquet Carneiro, Rio de Janeiro

|  | Total | Positive | Inconclusive | Undetected | p<sup>#</sup> |
| --- | --- | --- | --- | --- | --- |
| **Vaccine Status n (%)** |  |  |  |  |  |
| Influenza vaccine in 2019 | 750 (73.5) | 273 (73.5) | 61 (75.3) | 416 (73.2) | 0.925 |
| Influenza vaccine in 2020 | 428 (44.8) | 188 (51.6) | 36 (51.4) | 204 (39.1) | **0.001** |
| **Medical history n (%)** | **1,267** | **410 (32.4)** | **94 (7.4)** | **763 (60.2)** |  |
| Arterial Hypertension | 245 (19.3) | 94 (22.9) | 15 (15.9) | 136 (17.8) | 0.075 |
| Medicine drugs (in use) | 182 (14.3) | 80 (19.5) | 15 (15.9) | 87 (11.4) | **0.001** |
| Chronic Sinusitis | 117 (9.2) | 25 (6.1) | 8 (8.5) | 84 (11.0) | **0.021** |
| Asthma | 104 (8.2) | 26 (6.3) | 10 (10.6) | 68 (8.9) | 0.209 |
| Contraceptives (n=910) | 100 (10.9) | 31 (10.2) | 10 (13.7) | 59 (11.0) | 0.689 |
| Diabetes | 78 (6.1) | 29 (7.0) | 5 (5.3) | 44 (5.7) | 0.634 |
| Tobacco use | 73 (5.7) | 11 (2.6) | 4 (4.2) | 58 (7.6) | **0.002** |
| Obesity | 72 (5.6) | 24 (5.8) | 6 (6.3) | 42 (5.5) | 0.926 |
| Chronic Bronchitis | 64 (5.0) | 19 (4.6) | 7 (7.4) | 38 (4.9) | 0.527 |
| Alcohol addiction | 54 (4.2) | 22 (5.3) | 4 (4.2) | 28 (3.6) | 0.391 |
| Hypothyroidism | 46 (3.6) | 13 (3.1) | 6 (6.3) | 27 (3.5) | 0.316 |
| Cardiovascular disease | 25 (1.9) | 6 (1.4) | 2 (2.1) | 17 (2.2) | 0.664 |
| Tuberculosis | 14 (1.1) | 0 | 1 (1.0) | 13 (1.7) | **0.029** |
| Mental disease | 8 (.6) | 5 (1.2) | 0 | 3 (.3) | 0.170 |
| Neoplasia | 6 (.4) | 1 (.2) | 0 | 5 (.6) | 0.487 |
| Liver disease | 6 (.4) | 0 | 0 | 6 (.7) | 0.137 |
| Rheumatoid arthritis | 5 (.3) | 1 (0.2) | 0 | 4 (.5) | 0.626 |

**Table 3.** Age, signs and symptoms from COVID-19 nasopharyngeal swab individuals attended from March 18 to April 8, 2020 at Polyclinic Piquet Carneiro, Rio de Janeiro

|  | Total | Detected | Inconclusive | Undetected | P |
| --- | --- | --- | --- | --- | --- |
| Age (years) | **1,267** | 40.5 [34-49] (n=410) | 40 [34-50] (n=94) | 39 [33-50] (n=763) | 0.687 |
| Temperature | **1,010** | 36.8±0.8 (n=378) | 36.6±0.8 (n=86) | 36.5±0.8 (n=546) | 0.383 |
| Symptoms (days) m Q1-Q3 | **1,170** | 5 (4-7) (n=394) | 6 (4-7) (n=91) | 6 (4-8) (n=685) | **<0.0001<sup>a</sup>** |
| BMI | **926** | 28.3±5.5 (n=314) | 27.5±5.8 (n=69) | 27.6±5.4 (n=543) | 0.191 |
| Respiratory Frequency | **552** | 17.8±2.9 (n=176) | 17.6±2.9 (n=38) | 17.8±2.9 (n=338) | 0.854 |
| O2 saturation | **1,194** | 97.5±1.6 (n=396) | 97.7±1.4 (n=90) | 97.6±1.5 (n=708) | 0.557 |
| Cardiac Frequency | **851** | 86.1±14.6 (n=364) | 85.9±15.2 (n=71) | 84.9±14.8 (n=416) | 0.554 |
| Suspected Contact with Infected | **866 (72.8)** | 301 (77.6) | 69 (78.4) | 496 (69.6) | **0.008** |
| Contagious suspect days m Q1-Q3 | **708** | 7 [5.5-10] (n=256) | 8 [7-10] (n=555) | 8 [6-12] (n=397) | 0.060 |
| Confirmed Contact with Infected | **471 (40.6)** | 170 (43.8) | 43 (46.7) | 258 (38.0) | 0.081 |
| Contagious confirmed days m Q1-Q3 | **428** | 8 [6-10] (n=157) | 8 [7-10] (n=38) | 10 [7-13] (n=233) | 0.077 |
| **Symptoms n (%)** |  |  |  |  |  |
| Cough | **1,021 (80.5)** | 343 (83.6) | 77 (81.9) | 601 (78.7) | 0.123 |
| Headache | **880 (69.4)** | 310 (75.6) | 69 (73.4) | 501 (65.6) | **0.001** |
| Body ache | **800 (63.1)** | 318 (77.5) | 76 (80.8) | 406 (53.2) | **<0.0001<sup>a</sup>** |
| Fever | **711 (56.1)** | 291 (70.9) | 59 (62.7) | 361 (47.3) | **<0.0001<sup>a</sup>** |
| Sore throat | **666 (52.5)** | 193 (47.0) | 41 (43.6) | 432 (56.6) | **0.001** |
| Coryza | **650 (51.3)** | 193 (47.0) | 41 (43.6) | 416 (54.5) | **0.015** |
| Tiredness | **618 (48.7)** | 203 (49.5) | 58 (61.7) | 357 (46.7) | **0.022** |
| Sneeze | **527 (41.5)** | 164 (40.0) | 36 (38.3) | 327 (42.8) | 0.509 |
| Nasal congestion | **517 (40.8)** | 176 (42.9) | 40 (42.5) | 301 (39.4) | 0.481 |
| Prostration | **424 (33.4)** | 152 (37.0) | 34 (36.1) | 238 (31.1) | 0.106 |
| Respiratory difficulty | **346 (27.3)** | 94 (22.9) | 30 (31.9) | 222 (29.1) | **0.045** |
| Diarrhoea | **337 (26.6)** | 123 (30.0) | 21 (22.3) | 193 (25.2) | 0.137 |
| Anosmia or Hyposmia | **311 (24.5)** | 157 (38.2) | 31 (32.9) | 123 (16.1) | **<0.0001<sup>a</sup>** |
| Chill | **239 (18.8)** | 118 (28.7) | 22 (23.4) | 99 (12.9) | **<0.0001<sup>a</sup>** |
| Anxiety | **215 (16.9)** | 56 (13.6) | 24 (25.5) | 135 (17.6) | **0.015** |
| Nausea or Vomiting | **166 (13.1)** | 80 (19.5) | 18 (19.1) | 68 (8.9) | **<0.0001<sup>a</sup>** |
| Eye conjunctiva congestion | **147 (11.6)** | 41 (10.0) | 9 (9.5) | 97 (12.7) | 0.735 |
| Palpitation | **147 (11.6)** | 45 (10.9) | 13 (13.8) | 89 (11.6) | 0.313 |
| Irritability | **104 (8.2)** | 34 (8.2) | 9 (9.5) | 61 (7.9) | 0.868 |
| Abdominal pain | **102 (8.0)** | 49 (11.9) | 9 (9.5) | 44 (5.7) | **0.0009** |
| Sputum production | **66 (5.2)** | 32 (7.8) | 2 (2.1) | 32 (4.1) | **0.011** |
| Swallow difficulty | **44 (3.4)** | 18 (4.3) | 6 (6.3) | 20 (2.6) | 0.079 |
| Mental confusion | **25 (1.9)** | 7 (1.7) | 2 (2.1) | 16 (2.1) | 0.895 |
| Lymph node enlargement | **25 (1.9)** | 4 (0.9) | 1 (1.0) | 20 (2.6) | 0.124 |
| Skin rash | **14 (1.1)** | 7 (1.7) | 1 (1.0) | 6 (0.7) | 0.355 |

Cough, headache, body ache (including myalgia at rest or during minimal physical effort), fever, sore throat, coryza, and tiredness were common symptoms, present in more than a third of the patients in the cohort. These symptoms were common to both the undetected, inconclusive, and SARS-COV-2-positive groups. However, chills, anosmia or hyposmia, nausea or vomiting, and abdominal pain, although less frequent, were more common in individuals with the SARS-COV-2-positive results (p<0.001) than in patients with no detectable infection.

The respiratory and cardiac frequencies, temperature, O_2_ saturation, and laboratory results for this cohort are summarized in Table 4. CRP, LDH, AST, and ALT levels were all shown to be elevated in COVID-19 patients, while leukometry values were shown to differ between individuals regardless of the SARS-COV-2 SNF detection result.

**Table 4.** Laboratory results from COVID-19 nasopharyngeal swab individuals attended from March 18 to April 8. 2020 at Polyclinic Piquet Carneiro. Rio de Janeiro

|  | Positive | Inconclusive | Undetected | p |
| --- | --- | --- | --- | --- |
| N | 410 | 94 | 763 |  |
| CRP m (Q1-Q3) ng/L | 5.63 [2.32-13.49] | 4.52 [1.72-11.27] | 2.71 [1.12-7.37] | **<0.0001<sup>a,c</sup>** |
| ALT m (Q1-Q3) μkat/L | 0.37 [0.30-0.47] | 0.35 [0.27-0.48] | 0.30 [0.25-0.37] | **<0.0001<sup>a</sup>** |
| AST m (Q1-Q3) μkat/L | 0.37 [0.30-0.48] | 0.35 [0.22-0.63] | 0.30 [0.25-0.37] | **<0.0001<sup>a,c</sup>** |
| Creatinine μmol/L | 76.02±14.14 | 75.14±15.03 | 76.91±14.14 | 0.177 |
| LDH ukat/L | 3.05±7.62 | 3.10±8.77 | 2.95±7.65 | **0.031<sup>a</sup>** |
| CPK m (Q1-Q3) μkat/L | 1.27 [0.94-1.91] | 1.32 [0.89-1.95] | 1.39 [0.95-2.09] | 0.106 |
| N | 376 | 91 | 744 |  |
| Red cell $10^{12}/L$ | 4.8±0.4 | 4.8±0.5 | 4.8±0.5 | 0.834 |
| Haemoglobin g/L | 137±12 | 136±13 | 138±14 | 0.655 |
| Haematocrit % | 42±3.4 | 41.5±3.6 | 41.9±3.7 | 0.530 |
| MCV fL | 87.8±5.2 | 87.9±5.7 | 87.8±5.3 | 0.987 |
| MCH pg/cell | 28.7±1.8 | 28.8±2.3 | 28.8±2 | 0.609 |
| MCMH g/L | 326±10 | 328±10 | 328±10 | 0.064 |
| RDW % | 13.6±1.1 | 13.6±1.1 | 13.7±1.2 | 0.891 |
| Leukocytes m [Q1-Q3] $\times 10^9/L$ | 4,320 [3,620-5,360] | 4,850 [4,000-6,520] | 6,160 [4,985-7,445] | **<0.0001<sup>a,c</sup>** |
| Neutrophils (%) m [Q1-Q3] | 53 [45-63] | 55 [49-65] | 59 [52.5-65] | **<0.0001<sup>a,c</sup>** |
| Neutrophils (Abs) m [Q1-Q3] $\times 10^9/L$ | 2,379 [1,827-3,353] | 2,909 [2,161-4,116] | 3,780 [2,799-4,781] | **<0.0001<sup>a,c</sup>** |
| Lymphocytes (%) m [Q1-Q3] | 34 [27-42] | 34 [26-41] | 31 [26-38] | **<0.0001<sup>a,c</sup>** |
| Lymphocytes (Abs) m [Q1-Q3] $\times 10^9/L$ | 1,580 [1,278±2,014] | 1,920 [1,495±2,305] | 2,057 [1,663±2,473] | **<0.0001<sup>a,b,c</sup>** |
| Monocytes (%) m [Q1-Q3] | 17 [12±7] | 6 [12±8] | 6 [4±8] | **<0.0001<sup>a,b,c</sup>** |
| Monocytes (Abs) $\times 10^9/L$ | 473±196 | 470±167 | 503±192 | 0.181 |
| Eosinophils (%) m [Q1-Q3] | 0 [0-5.5] | 1 [0-2] | 1 [0-2] | **<0.0001<sup>a</sup>** |
| Eosinophils (Abs) m [Q1-Q3] $10^9/L$ | 0 [0-41] | 153 [0-44] | 215 [0-7] | **<0.0001<sup>a,b,c</sup>** |
| Basophils (%) | 0±0.2 | 0±0 | 0±0 | 0.329 |
| Platelets $10^9/L$ | 235±62 | 248±67 | 285±68 | **<0.0001<sup>a</sup>** |

Data expressed as mean ± SD. compared with ANOVA or by median and interquartile m [Q1-Q3] and compared with Kruskal-Wallis test and Tukey or Dunn tests for multiple comparison. **a-** Detected vs Undetected. **b-** Detected vs Inconclusive and **c-** Undetected vs Inconclusive (p<0.05).

When we applied the logistic regression, we observed that anosmia/hyposmia (OR: 2.38, 95%IC: 1.78-3.19, p < 0.0001), fever (OR: 2.13, 95%IC: 1.63-2.79, p < 0.0001), body pain (OR: 2.11, 95%IC: 1.59-2.82, p < 0.0001), and chills (OR: 1.70, 95%IC: 1.23-2.34, p = 0.001), in this specific order, were independent predictors for COVID-19. The other signs and symptoms did not make a significant contribution, when evaluated using the 95% confidence interval. The estimated probability of COVID-19 detection in patients presenting with these four signs and symptoms (fever, body pain, anosmia-hyposmia, and chills) was 72% when evaluated using a model adjusted for logistic regression.

When the laboratory test results (906 complete records) were evaluated using logistic regression leukocytes (p<0.0001), monocytes (p<0.0001), eosinophils (p<0.0001), CRP (p=0.001), and platelets (p=0.003), in this specific order, were significant independent predictors for COVID-19. The other laboratory variables did not make a significant contribution to predicting a SARS-CoV-2 positive sample at 95% confidence interval. For the final model, we considered the following variables: age, sex, onset of symptoms, education, laboratory results, and clinical signs and symptoms. Table 5 provides the final regression model, using 770 complete records to link specific data to SARS-COV-2 detection. We observed that leukopenia, monocytosis (%), eosinophil decrease %, onset of symptoms, increased CRP, presence of fever, decreased number of platelets, presence of anosmia/hyposmia, and body pain, in this specific order, were independent predictors for COVID-19.

**Table 5.** Logistic regression final with signs, symptoms and laboratory results independent predictors for detected COVID-19 in nasopharyngeal swab

| Variable | Coef±SE | Odds ratio | 95%CI | p |
| --- | --- | --- | --- | --- |
| Leukocytes | -0.0005±0.0001 | 0.99 | 0.99-0.99 | <0.0001 |
| Monocytes (%) | 1.1406±0.0334 | 1.15 | 1.07-1.22 | <0.0001 |
| Eosinophils (%) | -0.0074±0.0016 | 0.99 | 0.98-0.99 | <0.0001 |
| Symptoms onset | -0.1797±0.0406 | 0.84 | 0.77-0.90 | <0.0001 |
| C reactive protein | 0.0106±0.0042 | 1.01 | 1.00-1.02 | 0.012 |
| Fever | 0.5158±0.2227 | 1.68 | 1.08-2.59 | 0.020 |
| Platelets | -0.0052±0.0017 | 0.99 | 0.99-0.99 | 0.003 |
| Anosmia or Hyposmia | 0.5259±0.2438 | 1.69 | 1.05-2.73 | 0.031 |
| Body pain | 0.4590±0.2301 | 1.58 | 1.01-2.48 | 0.046 |

Coef±SE: coefficient ± standard error

### Symptoms and laboratory results for healthcare and non-healthcare personnel and their link to SARS-COV-2-positive SNF outcomes

CRP was significantly elevated in individuals with detectable SARS-COV-2, as shown in table 4 (reference value up to 8 mg/L), with this increase (93.5%) being more prominent in the HCP group.

LDH was slightly elevated, although within the reference range (2.25-3.75 µkat/L), in SARS-COV-2-positive samples. An increase of 22% was observed in transaminases in SARS-CoV-2-positive samples from non-healthcare personnel (Non-HCP) compared to a 28% increase in HCP; additionally, this increase remained within the reference range (ALT results up to 0.55 µkat/L in women and 0.68 µkat/L in men and AST results up to 0.53 µkat/L in women and 0.63 µkat/L in men are considered normal).

The mean total for the leukocytes decreased by 29%-32% in the presence of infection and was connected with reduced absolute neutrophil, lymphocyte, and eosinophil counts. However, there was a slight increase in the absolute monocyte counts in positive samples which coincided with a minor increase in the relative lymphocyte counts. Additionally, SARS-CoV-2-positive samples exhibited a 15%-22% reduction in their platelet count as well.

### Detection of SARS-COV-2 in healthcare professionals and non-healthcare personnel as a function of time

There was a correlation between the frequency of SARS-COV-2 detection and the week of testing with the HCP incidence curve appearing to be parallel with the Non-HCP incidence curve (Figure 1).

**Figure 1.**
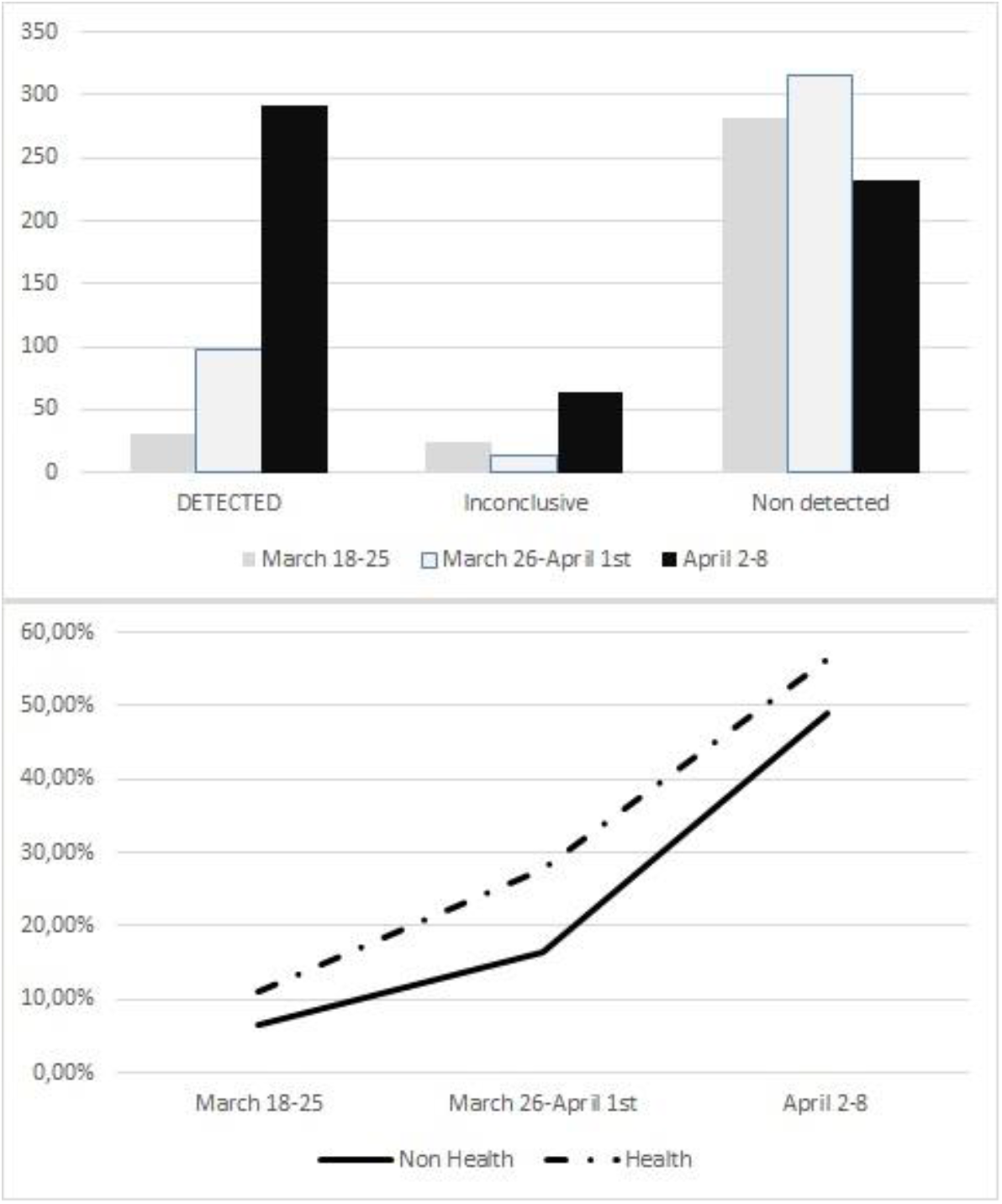
COVID-19 results during the time interval (weeks). a) Frequency of detected. inconclusive and undetected results. b) Non Healthcare personnel *vs* Healthcare professionals detected COVID-19 frequency curves.

## DISCUSSION

Although COVID-19 is known for its wide variety of clinical symptoms, we were able to identify some differences between casual and continuous reports. Findings of fever, anosmia or hyposmia, body pain, and chills were found to be linked to a 72% probability for a positive diagnosis, making these the most reliable clinical symptoms of infection.

### Characteristics of patients

While this initiative was initially designed to support HCP at our institution, the dire need of expansive clinical support and diagnosis for COVID-19 meant that, from week two, we included patients from other public institutions and patients frequently treated at our facility in our testing cohort. This meant that while most of the individuals (80%) evaluated and tested at our site were HCP, 20% of the study cohort comprised Non-HCP patients.

The HCP group predominantly comprised self-identified white professionals (55.5%) with at least 12 years of study (80.1%) while only 42.2% of the individuals in the non-HCP group identified as white. The Non-HCP group had a higher frequency of obesity (11.1% vs 4.4%) compared to that in the HCP group which was reflected in the BMI values (Non-HCP: 29.2±6.0 vs HCP: 27.4±5.2). Additionally, smoking was also more frequent in the Non-HCP group, and there were also differences in adherence to the influenza vaccination scheduled between these two groups.

As of April 8, 2020, the deadline for data collection, Brazil had reported a total of 15,927 confirmed cases of COVID-19.[3] The State of Rio de Janeiro had 1,938 cases with this study representing 21.6% of all the cases reported in this state, with the majority of these cases being reported in HCP.

### SARS-COV-2 nasopharyngeal swab results

Here, we noted that detection of the SARS-COV-2 RNA in NPS is associated with the onset of COVID-19 symptoms. This data is consistent with previous studies which identifies that the virus is detectable within the first eight days of symptoms [4,5] and reinforces the need for a differentiated strategy for determination of eligibility for NPS SARS-COV-2 testing, especially where the health system is constrained in terms of both human resources and the availability of test kits. Early recognition of COVID-19 is critical for the isolation of infected persons and the mitigation of community spread.[6] The ability to rule-out COVID-19 earlier enables more effective public health containment measures and frees up resources by reducing isolation time in suspected cases. Our study suggests that there is sufficient viral load to allow a positive NPS result within 3-4 days of infection, and this may significantly reduce isolation times, thus freeing up many valuable resources.

Headache, body ache, fever, anosmia or hyposmia, and chills were common symptoms and were more frequent among SARS-COV-2 NPS-positive patients than in SARS-COV-2 NPS-negative patients. These are well-known signs and symptoms of various viral infections and COVID testing should be reliant on a combination of these symptoms and a comprehensive understanding of the patients’ local, regional, and travel history especially in resource-constrained conditions. HCP should treat any patient with these symptoms and a high likelihood of exposure as SARS-CoV-2-positive even in the absence of an appropriate test. Zhang et al.[7] observed fever (91.7%), cough (75.0%), fatigue (75.0%), and gastrointestinal symptoms (39.6%) in their COVID-19 cohort. In a meta-analysis, Li and colleagues [5] reported that fever (88.5%), cough (68.6%), myalgia or fatigue (35.8%), expectoration (28.2%), and dyspnea (21.9%) were all primary indicators of COVID-19. In this study, we also investigated if the combination of signs and symptoms could help in developing a decision-making rationale for ordering laboratory tests. Curiously, an increase in the frequency of anxiety and “difficulty in swallowing” was observed among those patients with inconclusive SARS-COV-2 NPS results. Psychological aspects and global mental health have already been the subject of a number of studies, as the mental toll of the patient becomes more evident.[8-10] In China, more than 70% of the patients experience moderate to high levels of psychological symptoms, specifically elevated scores for obsessive compulsion, interpersonal sensitivity, phobic anxiety, and psychoticism.[11] Our results did not corroborate those of Zhang and colleagues [12], who showed a higher prevalence of insomnia (38.4% vs. 30.5%, p < 0.01) and anxiety (13.0%vs. 8.5%, p < 0.01) in Non-HCP (n = 1,255) patients, while we found no differences in the prevalence of anxiety between HCP and Non-HCP in our study. This may be due to the methodology used, as this study was not designed to compare these two groups. It is not possible to further characterize the work and professional activity of our participants but it is worth noting that there have been other studies that have identified differences in anxiety and stress among medical and nursing staff [13].

Anosmia is a frequent symptom [14] of COVID-19 and is often the only indicator in asymptomatic carriers.[15] This symptom is transient and it has been suggested that this anosmia is linked to the destruction of the epithelial cells in the nose. These cells have a high expression of angiotensin converting enzyme 2, the receptor for SARS-CoV-2, which allows the virus to penetrate the system.[16]

Dyspnea is a well-characterized symptom of COVID-19 and has been shown to be closely related to the severity of the disease and mortality rates.[17,18] Our study did not evaluate disease severity, but rather focused on identification of clinical and laboratory data that could be used to predict COVID-19 infection in HCP, in an effort to allow for their rapid isolation to prevent wide spread infections among the hospital staff. Our results showed that anosmia / hyposmia (p < 0.0001), fever (p < 0,0001), body pain (p < 0.0001), and chills(p = 0.001), in this specific order, were independent predictors for SARS-COV-2 NSF detection. Moreover, the combination of these symptoms has a prognostic capacity of around 72% for SARS-COV-2. This information should guide healthcare workers in determining the course of evaluations including laboratory testing, while allowing for the fact that the COVID-19 pandemic may contribute to revisions in the State and Municipal health authority guidelines for clinical testing and isolation.

The laboratory results indicate that the total leukocytes, % of monocytes, eosinophils, RCP, and platelet counts were all independent predictors of COVID-19. These factors were also identified in several previous COVID-19 studies.[5 19 20] In this study, we were able to combine signs, symptoms, and laboratory results in a logistic regression analysis that identified various independent predictor values that could be used to supplement RT-PCR testing in resource scarce environments. Leukocyte counts of under 5,400/µL, relative monocytosis (>9%), eosinopenia, and elevated RCP within the first seven days from the onset of fever, anosmia / hyposmia, and body pain were the main independent variables associated with positive RT-PCR results. Although platelets also appeared to be an independent predictor, the range for this value remained within the clinical reference values and would need to be compared with previous results from the same patient to be of a significant value. Additionally, the cut-off points for leukocyte counts and percentage of monocytes may also guide the interpretation of the complete blood cell count during the first days of infection. Additionally, these results may also be used during follow-up for hospitalized patients when AST, ALT, gamma-GT, LDH, and alpha-HBDH are all markedly altered [21].

The risk factors reported here may be used in future studies to evaluate associated medical interventions, hospitalization, and mortality rates. This cohort should also be expanded and new longitudinal evaluations should be undertaken to better understand the condition of these patients in a post pandemic future. Nonetheless, we were able to confirm a greater risk for infection in HCP. It is well known that HCP are subject to far greater continuous exposure to COVID-19 than the general public, thus increasing their infection rates. Noteworthy, this screening started at the beginning of the epidemic in Brazil and that the number of detected cases in this study retained a linear scale, suggesting that early infections with COVID-19 were more prevalent among HCP and for this reason close monitoring of this population might be crucial to reducing the burden on the healthcare system.

This study has several limitations associated with the fact that it is a cross-sectional, ongoing study, with an increasing number of patients. Although limited in number, data here may support other studies so that in the long term a better meta-analysed prediction system can be developed. The HCP population is predominantly made up of younger people and this may cause differences in the observations of this study versus studies conducted in older populations.[22 23] Currently, laboratory-based RT-PCR is the recommended test for diagnoses of acute cases to ensure patients can be identified and isolated and to facilitate the public health response, as reviewed by Venter and Richter {Venter, 2020 #1796}. On the other hand, The false-negative rate has important ramifications for gaining accurate clinical and epidemiological data and false-negative results may lead to misdiagnoses in both patients and HCP, with increased risk of infection transmission {Payne, 2020 #1795}. So, combining routine laboratory test that can detect both acute phase proteins and specific altered leukocyte profile corresponding to COVID-19 infection could add parameters to more efficient control of suspected individuals. Nonetheless, the rapid development of point-of-care molecular or antigen tests are already a reality {Lippi, 2020 #1468}

Up to May. 2020, the clinical diagnosis and medical intervention for COVID-19 has been restricted in HCP servicing the military, civil, and federal police reaching 1,626 confirmed cases.

In conclusion, positive SARS-COV-2 NPS assays were associated with fever, anosmia or hyposmia, body pain, and chills, and patients were all positive within the first 3-8 days after symptom onset. Elevated CRP, leukocyte counts under 5,400 /µL, and relative monocytosis (>9%) were all common among patients with COVID-19. These parameters may help to identify and isolate probable SARS-CoV-2 infections in the absence of RT-PCR tests.

## Data Availability

Data is available upon request.

## Key message

- Healthcare professionals (HCP) working at the frontline are among the groups most affected by the COVID-19 pandemic. We investigated the clinical features of 1,297 individuals, mostly (80%) HCP, who tested positive for the SARS-CoV-2 (COVID-19) during the period immediately following the pandemic declaration in Brazil (March 19 to April 8, 2020).
- Signs and symptom data and blood samples were collected for all individuals who were tested for SARS-CoV-2 by RT-PCR assays in nasopharyngeal swab (NPS).
- CRP, LDH, AST, and ALT were elevated among COVID-19-infected individuals, and leukometry differed between individuals within the different SARS-COV-2 NPS detection results. Anosmia/hyposmia, fever, body pain, and chills, in this specific order, were independent predictors for COVID-19. The estimated probability of COVID-19 detection when all four of these signs and symptoms were present was 72%. There was a trend of increasing number of SARS-CoV-2-positive test results in later weeks of analysis especially in HCP samples.
- Here, we identified that the significant clinical features of SARS-CoV-2 in Brazil included anosmia, fever, chills, body pain, elevated CRP, leukocytes under 5,400 x 10^9^/L, and relative monocytosis (>9%). These variables may help, in the absence of RT-PCR tests, to monitor the COVID-19 infection rate during a pandemic.

## Authors contribution

Study conception and design: L.C. Porto, R. Rufino, C.H. Costa, A. S. Nunes and I. Bouzas

Interview, Consultation and Patient orientation and NPS and blood sampling: A. S. Nunes, T. C. Ferreira, A.C. Faria, F.H. Silva, C. A. Duarte, S. Vilas Boas

Laboratory testing and interpretation: D.A. Secco, A. M. G. Santos, R. S. Menezes, V. M. A. Souza, V. M. Porto, J. R. Mota, A. P. Villela-Silva, M. V. Castro, J. S. Nogueira, F. M. Vasconcelos, A. D. Silva, C. V. Lima and N. B. Brazão, I. A.A. Brito, S. B. Costa, R. M. R. Lemes, J. I. O. dePaula, G. L. Bongionanni, O. C. L. Bezerra, V. V. Andrade-Neto, L. Morette.

Electronic Data Acquisition: S.M. Freire, I. Bouzas, R. Souza.

Logistic, Communication and Supply services: E. P. Bittencourt, A. V. Loureiro, R. S. Barboza, J. A L. Sanches, D. P. Felizardo, A. V. Rodrigues, P. A. Franco, R. Miranda

Analysis of data and figures: R. A. G. Martins, I. Bouzas, R. Rufino, C. H. Costa and L.C. Porto

Interpretation of data: I. Bouzas, R. Rufino, C. H. Costa and L.C. Porto

Drafting of manuscript: L. C. Porto, R. Rufino and C. H. Costa.

Critical revision: C. H. Costa and I. Bouzas

All authors approved the final manuscript.

## Acknowledgments

We thank all team at the front line for reception of patients, security and cleaning services. Undergraduate, interns and residents acting as volunteers were also deeply acknowledge. We also thank Carlos Nunes, Andrea Benazzi, Karina Goulart, Tainah Lima, Miguel Garcia, Breno Sampaio and Marcos Alcoforado for data entry and Mário João Junior for electronic entry data development. We would like to thank Ana Clara Perrone and Editage (www.editage.com) for English language editing.

